# Primary healthcare facilities readiness for animal-related injuries: Availability, stock-Outs, and affordability of essential commodities in rural public health facilities, Tanzania

**DOI:** 10.64898/2026.08.03.26359542

**Authors:** Manase Kilonzi, Joseph Matobo Thobias, Gimbo Hyuha, Paul Makoye Malaba, Beatrice Aiko, George Kiwango, Alphonce Ignace Marealle, Nathanael Sirili

## Abstract

**Background:** In Tanzania, reported cases for animal-related injuries is approximated to be 200-40,000, where most of deaths and complications from these cases are entirely preventable when safe, affordable and effective medications are available and administered in a timely manner.

**Methodology/Principal findings:** This study analyzed the availability, stockouts, and affordability of essential animal-related injury health commodities at primary healthcare (PHC) facilities in rural settings in Tanzania. A retrospective cross-sectional study was conducted in February 2024, using health facility data covering the period from 2019 to 2023. Availability, stockouts, and price of the selected 33 health commodities were extracted from hospital pharmaceutical ledgers at 29 selected public PHC facilities in Mkinga District, Tanga region comprising 26 dispensaries and three health centers. Affordability was calculated using the wage of the lowest-paid government worker per day approach. Overall availability remained below the WHO target of > 80% and dispensaries were more affected than health centers. Notable incremental availability was observed in 2023; however, differences were observed between dispensaries and health centers; snake antivenom (34.6% vs 100%), antirabies (23.1% vs 66.7%), adrenaline injection (73.1% vs 100%), and dexamethasone injection (0% vs 66.7%). Across the five years, dispensaries had more stockouts between 2019 and 2021, reporting up to a 53.8% stockout rate for chlorpheniramine tablets, with the longest stockout period of 218 days for an IV giving set in 2019. The health centers had higher stockouts between 2021 and 2022, reporting up to a 66.7% stockout rate for hydrocortisone injection in 2021 and the longest stockout period of 102 days for dexamethasone injection in 2020. Using the affordability threshold of 7% of daily income for healthcare expenditure, nearly all assessed commodities were classified as unaffordable.

**Conclusions/Significance:** Essential commodities for managing animal-related injuries remain inadequately available, frequently out of stock, and largely unaffordable in rural PHC facilities in Tanzania, particularly anti-rabies biologicals and snake antivenom. Strengthening surveillance, forecasting, procurement, financing, and supply chain systems, alongside expanding financial risk protection through health insurance and targeted subsidies, is essential to improve equitable access to life-saving commodities.

**Author Summary:** Most of deaths and complications from animal-related cases are entirely preventable when safe, affordable and effective medications are available and administered in a timely manner. In this study we analyzed availability, stockout and affordability of the of essential animal-related injury health commodities at primary healthcare facilities in rural settings in Tanzania. Essential health commodities for managing animal-related injuries were available below the WHO-recommended target, with anti-rabies biologicals and snake antivenoms showing the poorest availability. Frequent and prolonged stockouts, particularly in dispensaries, substantially limited continuous access to life-saving medicines and medical supplies for animal-related injuries. Nearly all assessed commodities were unaffordable using the lowest-paid government worker benchmark, highlighting the need to strengthen supply chain systems, forecasting, and sustainable financing to improve equitable access.

## Introduction

Globally, animal-related injuries are a public health challenge, with nearly half of the world’s population experiencing at least one such injury during their lifetime [1,2]. Worldwide, an estimated two to five million people are at risk, resulting in 3,000-137,880 mortalities and more than 413,000 case-morbidities annually [3–5]. Sub-Saharan Africa (SSA) experiences nearly one million cases each year, including over 100,000 envenomation and an estimated 10,000-30,000 deaths [6,7]. In Tanzania, reported cases have risen markedly in recent decades, approximately 200-40,000 cases reported over decades [5–10]. According to the WHO, most deaths and severe complications from animal-related injuries are preventable through timely access to safe and effective antivenoms, antibiotics, and other essential medical interventions [5].

Access to safe, effective, timely, affordable, and quality essential medicines has been included in the United Nations’ Sustainable Development Goals (SDGs) as an important element towards universal health coverage (UHC) [12–14]. The WHO recommends a minimum availability target of 80% for essential medicines across all member states [7]. However, more than one-third of the global population lacks reliable access to essential medicine, with nearly half of the population in SSA and Asia being affected [16–19]. Achieving the WHO target remains a major challenge in many SSA countries due to multiple factors, including poverty, high cost of medicine and medical devices, poor inventory management, poor forecasting, limited implementation of the UHC drug act, importation of low-quality medical products, and limited local pharmaceutical industries [17,20]. Consequently, inadequate access to essential medicines contributes to catastrophic health expenditure at the individual, family, community, and national level, further perpetuating a vicious cycle of poverty.

The situation is particularly pronounced for essential medicines and medical products used in the management of animal-related injuries, which fall within the broader category of neglected tropical diseases (NTD). For instance, the availability of snake antivenom in urban Rwanda was below five percent among the surveyed facilities [10]. Medications like snake antivenom, prednisolone, and hydrocortisone, which are key in the management of a vast selection of animal-related injuries, were often unaffordable, requiring patients or their relatives to work between one and ten days, based on minimum wage estimates, to cover a single day’s treatment costs [17,19]. Similarly, in rural and urban Kenya, the overall availability of the snakebite commodities was below 50%, and stockouts of essential medicines for snakebite management were approximately 18% and 11% in public and private facilities, respectively, over one year [11].

In Tanzania, efforts have been made over recent decades to strengthen the health system, particularly in the building of healthcare infrastructure, training and capacity building for Human Resources for Health (HRH), and procurement of essential medicines and medical devices [21–23]. Recent reports show that the availability of essential medicines and the presence of emergency care services (EMS) at regional referral, zonal-referral, specialized, and national hospitals are satisfactory [24,25]. Besides initiatives to strengthen healthcare systems at PHC levels, which serve as the first point of care for over 70% of people, with 97 private and public district hospitals having formalized emergency departments, there is an ongoing project piloting the establishment of emergency services at health centers and dispensaries [14].

Although the availability and affordability of essential medicines and medical products have been explored in many LMICs, studies that are specifically assessing medicines and medical products for animal-related injuries are limited, particularly in rural settings where the burden is enormous. Given that animal-related injuries are recognized among NTDs, understanding access to essential commodities for their management is critical. This study, therefore, analyzed the availability, stockouts, and affordability of essential animal-related commodities in PHC facilities in rural Tanzania across five years. The findings contribute to the limited evidence base on access to essential animal-related injury commodities in rural settings in developing countries and provide important information for policymakers as efforts to strengthen PHC systems and achieve UHC continue in Tanzania and other similar settings.

## Methods

### Study design

This retrospective descriptive cross-sectional study was conducted in February 2024 in rural public PHC facilities in Mkinga District, Tanga Region, Tanzania. We retrospectively analyzed health facility data covering a period from January 2019 to December 2023. The study was nested in a broader study that was conducted to review incidents, clinical characteristics, and management of animal-related injuries in the locality [5,26]. The present paper focuses on reviewing the availability, stockout, and affordability of essential health commodities used in managing animal-related injuries between 2019 and 2023 [5,26].

### Study settings

The Tanzania health system is a pyramid structure, from the bottom (primary level) to the top (tertiary level). It is pluralistic in nature, comprising public, private, faith-based, non-profit, and both modern and traditional medicine actors delivering healthcare services. Dispensaries and health centers are the lowest tier of health facilities [16]. This study was conducted in PHC-level facilities in Mkinga District, located in the Tanga Region, along the Indian Ocean. Mkinga is a rural district with a population of 146,802 per the 2022 census, covering 2,712 km^2^ [17]. The district was selected for this study because previous research has reported a high burden of animal-related injuries [28]. By February 2024, the district had 42 public functional healthcare facilities, including 3 health centers and 39 dispensaries.

### Sampling of facilities

Of the 42 functional public PHC facilities in Mkinga District, 29 were purposively selected for inclusion in the study. Facilities were eligible if they had been operational continuously for at least the preceding three years.

### Data collection procedure

A data extraction tool was developed based on the WHO/ Health Action International (HAI) structure to assess the availability, prices, and affordability of medicines at healthcare facilities. The data extraction tool requires that both core and supplementary list of medicines based on local needs and disease burden of a particular country should be included. In this study, 33 core essential medicines and medical devices from the Standard Treatment Guidelines and National Essential Medicines List (STG-NEML) for Tanzania Mainland [18] were included.. Out of 33 selected commodities, 21 were drugs/medicines, of which 14 items were injectables and intravenous (IV) fluids, and 7 items were oral/topical medications. Twelve were medical devices/tests: eleven were medical devices for emergency care, and one was a diagnostic test.

For each medicine and medical device, the following were collected for each year (2019 – 2023): availability, lowest unit price, stockout, and longest stockout period. All this information was extracted physically from the respective facility’s pharmaceutical main store ledgers.

Five research assistants (Ras) were recruited from the Mkinga district. The recruited RAs were healthcare providers employed in Mkinga district, with diplomas and certificates in medicine, pharmacy, and nursing. Recruited RAs were actively involved in ordering and receiving medicines and medical devices in their facility; therefore, they were aware of the use of ledgers. The Mkinga District Medical Officer was consulted during the selection of the RAs. A one-day workshop was conducted to orient RAs on the importance of the study and how to fill out the data extraction tool. On the day of the workshop, each RA was asked to bring one completed ledger from their facility for practice. Thereafter, the district was divided into five zones (each with 5-6 facilities), and each RA was assigned one zone.

### Data analysis

Data were entered, cleaned, and analyzed using Microsoft Excel. Findings are summarized using frequency and percentages.

### Medicine and medical device availability

Availability of each commodity was assessed at the facility level and defined as the presence of the commodity during the assessment period of 2019-2023. Availability was calculated as the proportion of health facilities with the commodity available divided by the total number of facilities assessed at each category of PHC (dispensaries and health centers), multiplied by 100. For dispensaries, percentages were calculated using 26 facilities as the denominator, while for health centers, percentages were calculated using 3 facilities as the denominator. The availability was then defined according to the WHO scale of < 30% was considered very low, 30-49% low, 50-80% fairly high, and > 80% as high.

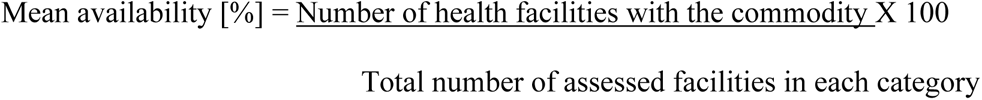

### Medicine and medical device stockout

A stockout was defined as the absence of a commodity at a health facility in each year. The stockout rate was calculated as the number of health facilities experiencing at least one stockout episode of a commodity divided by the total number of assessed facilities within each facility category [dispensary and health center], multiplied by 100. The longest stockout period was defined as the number of days a commodity remained unavailable in a facility during a given reporting year. The mean longest stockout duration was then calculated by averaging the longest stockout durations among facilities per category reporting stockouts.

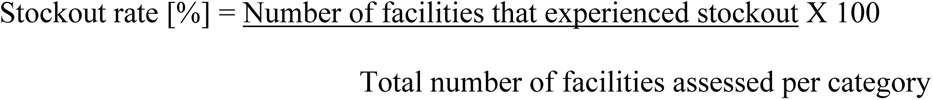

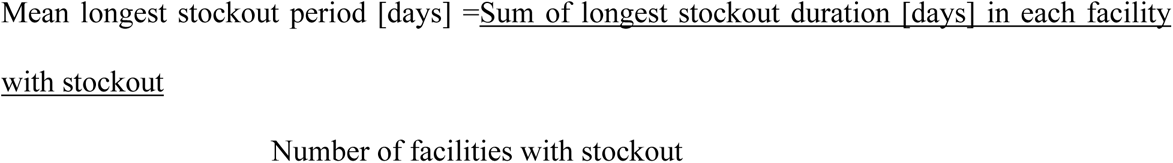

### Medicine and medical device affordability

According to the WHO/HAI methodology, we determined the affordability of animal-related commodities by the number of days of income required for an LPGW to cover the cost of the commodity for one full course of treatment for an animal-related injury. The commodity that costs the equivalent of one day’s wage of the LPGW to purchase a full course of treatment for an animal-related injury was generally considered affordable, and treatment that costs more than one day’s wage of the LPGW was considered unaffordable. In Tanzania, the lowest-paid unskilled government worker earned 3,077 TZS (∼ 1.19USD) per day [19]. Affordability was calculated based on Low Government Wage (LGW) per day = (3,077TZS) ∼ 1.19US

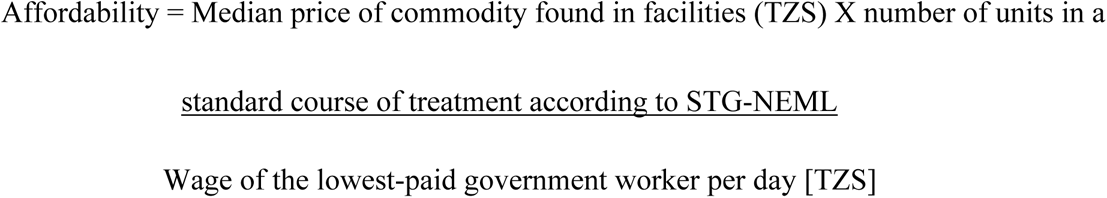

### Ethical Approval

Ethical approval for this study was obtained from the Muhimbili University of Health and Allied Sciences (MUHAS) Research and Ethics Committee (REC) (Approval No. MUHAS-REC-07-2023-1813). We obtained permission to collect data from the Ministry, the Tanga region, and the Mkinga district administration. All procedures followed the Declaration of Helsinki. All collected data were securely stored and accessible only to the principal investigator and authorized research team members.

## Results

### Availability of animal-related injuries commodities

Overall, the availability of assessed commodities remained below the WHO target of 80%. Anti-rabies vaccine availability was particularly low in dispensaries, ranging from 3.8% to 23.1% between 2019 and 2023, while health centers showed modest improvement from 33.3% to 66.7% over the same period. Snake antivenom availability was also critically low in dispensaries (3.8%–34.6%), but increased in health centers from 33.3% during 2019–2021 to 100% from 2022 onwards. In contrast, tetanus toxoid, hydrocortisone injection, prednisolone tablets, normal saline, Ringer lactate, and essential emergency supplies (Ambu bags, IV cannulas, and IV giving sets) were consistently available at ≥80% across most facilities. The availability of dextrose normal saline in dispensaries improved from 69.2% in 2019 to 84.6% in 2022/2023. However, dexamethasone injection remained scarce, particularly in dispensaries (0.0%–3.8%) despite moderate availability in health centers (33.3%–66.7%). Urine dipsticks also remained inadequately available, with availability below 43% in dispensaries and 67% in health centers throughout the study period. Advanced airway equipment, including endotracheal tubes and laryngeal mask airways, remained poorly available, particularly in dispensaries [<4%], while availability in health centers did not exceed 67% (Table 1).

**Table 1.** Availability of animal-related injury commodities in visited Public Health Facilities in Mkinga District, Dispensary (n=26) and Health center (HC) (n=03).

| Commodity | 2019 |  | 2020 |  | 2021 |  | 2022 |  | 2023 |  |
| --- | --- | --- | --- | --- | --- | --- | --- | --- | --- | --- |
|  | Dispen<br>sary<br>n (%) | HC<br>n (%) | Dispensar<br>y<br>n (%) | HC<br>n (%) | Dispensar<br>y<br>n (%) | HC<br>n (%) | Dispensar<br>y<br>n (%) | HC<br>n (%) | Dispensar<br>y<br>n (%) | HC<br>n (%) |
| **Antidote** |  |  |  |  |  |  |  |  |  |  |
| Anti-rabies<br>injection | 1 (3.8) | 1<br>(33.3) | 1 (3.8) | 1<br>(33.3) | 1 (3.8) | 1 (33.3) | 2 (7.7) | 2 (66.7) | 6 (23.1) | 2<br>(66.7) |
| Tetanus toxoid | 23<br>(88.5) | 3<br>(100) | 23 (88.5) | 3<br>(100) | 24 (92.3) | 3 (100) | 23 (88.5) | 3 (100) | 23 (88.5) | 2<br>(66.7) |
| Snake antivenom | 1 (3.8) | 1<br>(33.3) | 1 (3.8) | 1<br>(33.3) | 1 (3.8) | 1 (33.3) | 2 (7.7) | 3 (100) | 9 (34.6) | 3<br>(100) |
| Adrenaline<br>injection | 16<br>(61.5) | 3<br>(100) | 12 (46.2) | 2<br>(66.7) | 12 (46.2) | 2 (66.7) | 15 (57.7) | 3 (100) | 19 (73.1) | 3<br>(100) |
| **Corticosteroids** |  |  |  |  |  |  |  |  |  |  |
| Dexamethasone<br>injection | 0 (0.0) | 2<br>(66.7) | 1 (3.8) | 2<br>(66.7) | 1 (3.8) | 1 (33.3) | 1 (3.8) | 2 (66.7) | 0 (0.0) | 2<br>(66.7) |
| Hydrocortisone<br>injection | 21<br>(80.8) | 3<br>(100) | 19 (73.1) | 3<br>(100) | 20 (76.9) | 3 (100) | 22 (84.6) | 3 (100) | 25 (96.2) | 3<br>(100) |
| Prednisolone | 21 | 3 | 22 (84.6) | 3 | 22 (84.6) | 2 (66.7) | 23 (88.5) | 3 (100) | 25 (96.2) | 3 |
| tablets | (80.8) | (100) |  | (100) |  |  |  |  |  | (100) |
| **Antihistamine** |  |  |  |  |  |  |  |  |  |  |
| Cetirizine tablets | 15<br>(57.7) | 3<br>(100) | 15 (57.7) | 2<br>(66.7) | 12 (46.2) | 3 (100) | 20 (76.9) | 3 (100) | 23 (88.5) | 3<br>(100) |
| Promethazine<br>injection | 11<br>(42.3) | 3<br>(100) | 13 (50.0) | 3<br>(100) | 11 (42.3) | 1 (33.3) | 11 (42.3) | 2 (66.7) | 14 (53.8) | 3<br>(100) |
| Chlorpheniramine<br>tablets/injections<br>* | 21<br>(80.7) | 3<br>(100.0) | 23 (88.4) | 3<br>(100.0) | 24 (92.3) | 3 (100.0) | 25 (96.2) | 3 (100.0) | 26 (100.0) | 3<br>(100.0) |
| Promethazine<br>tablets | 14<br>(53.8) | 3<br>(100) | 14 (53.8) | 2<br>(7.7) | 16 (61.5) | 3 (100) | 18 (69.2) | 2 (66.7) | 20 (76.9) | 3<br>(100) |
| **Analgesia** |  |  |  |  |  |  |  |  |  |  |
| Paracetamol<br>tablets | 24<br>(92.3) | 3<br>(100) | 25 (96.2) | 3<br>(100) | 23 (88.5) | 3 (100) | 26 (100) | 3 (100) | 26 (100) | 3<br>(100) |
| Paracetamol<br>injection | 0 (0.0) | 1<br>(33.3) | 0 (0.0) | 1<br>(33.3) | 0 (0.0) | 1 (33.3) | 0 (0.0) | 0 (0.0) | 0 (0.0) | 1<br>(33.3) |
| Ibuprofen<br>tablets/syrup | 6<br>(23.1) | 2<br>(66.7) | 7 (26.9) | 2<br>(66.7) | 7 (26.9) | 3 (100) | 10 (38.5) | 2 (66.7) | 15 (57.7) | 2<br>(66.7) |
| Diclofenac<br>injection | 19<br>(73.1) | 3<br>(100) | 23 (88.5) | 3<br>(100) | 22 (84.6) | 3 (100) | 25 (96.2) | 2 (66.7) | 25 (96.2) | 3<br>(100) |
| Acetylsalicylic acid tablets | 4<br>(15.4) | 2<br>(66.7) | 3 (11.5) | 2<br>(66.7) | 5 (19.2) | 2 (66.7) | 4 (15.4) | 2 (66.7) | 6 (23.1) | 2<br>(66.7) |
| **Fluids for Infusion** |  |  |  |  |  |  |  |  |  |  |
| Dextrose Normal Saline | 15<br>(57.7) | 3<br>(100) | 15 (57.7) | 3<br>(100) | 15 (57.7) | 2 (66.7) | 18 (69.2) | 3 (100) | 19 (73.1) | 3<br>(100) |
| Normal saline | 22<br>(84.6) | 3<br>(100) | 21 (80.8) | 3<br>(100) | 21 (80.8) | 2 (66.7) | 24 (92.3) | 3 (100) | 24 (92.3) | 3<br>(100) |
| Ringer lactate | 22<br>(84.6) | 3<br>(100) | 25 (96.2) | 3<br>(100) | 22 (84.6) | 3 (100) | 23 (88.5) | 3 (100) | 25 (96.2) | 3<br>(100) |
| **Local anesthesia** |  |  |  |  |  |  |  |  |  |  |
| 1% Lignocaine injection | 22<br>(84.6) | 3<br>(100) | 23 (88.5) | 3<br>(100) | 23 (88.5) | 3 (100) | 23 (88.5) | 3 (100) | 24 (92.3) | 3<br>(100) |
| **Laboratory test** |  |  |  |  |  |  |  |  |  |  |
| Urine dipsticks | 8 (3.8) | 2<br>(66.7) | 8 (3.8) | 2<br>(66.7) | 8 (30.8) | 2 (66.7) | 11 (42.3) | 2 (66.7) | 11 (42.3) | 2<br>(66.7) |
| **Lifesaving Equipment** |  |  |  |  |  |  |  |  |  |  |
| Ambu bag<br>(Manual resuscitator) | 22<br>(84.6) | 3<br>(100) | 21 (80.8) | 3<br>(100) | 21 (80.8) | 3 (100) | 21 (80.8) | 3 (100) | 21 (80.8) | 3<br>(100) |
| Endotracheal | 1 (3.8) | 2 | 1 (3.8) | 2 | 1 (3.8) | 2 (66.7) | 1 (3.8) | 2 (66.7) | 1 (3.8) | 2 |
| intubation tube |  | (66.7) |  | (66.7) |  |  |  |  |  | (66.7) |
| IV cannula | 25<br>(96.2) | 3<br>(100) | 25 (96.2) | 3<br>(100) | 24 (92.3) | 3 (100) | 26 (100) | 3 (100) | 26 (100) | 3<br>(100) |
| IV giving set | 24<br>(92.3) | 3<br>(100) | 24 (92.3) | 3<br>(100) | 25 (96.1) | 3 (100) | 26 (100) | 3 (100) | 26 (100) | 3<br>(100) |
| Laryngeal mask<br>airway (LMA) | 1<br>((3.8) | 1<br>(33.3) | 1 (3.8) | 1<br>(33.3) | 1 (3.8) | 1 (33.3) | 1 (3.8) | 1 (33.3) | 1 (3.8) | 1<br>(33.3) |
| Nasal gastric<br>tube | 1<br>((3.8) | 1<br>(33.3) | 1 (3.8) | 1<br>(33.3) | 1 (3.8) | 1 (33.3) | 1 (3.8) | 1 (33.3) | 1 (3.8) | 1<br>(33.3) |
| Nasal prongs | 1<br>((3.8) | 1<br>(33.3) | 1 (3.8) | 1<br>(33.3) | 1 (3.8) | 1 (33.3) | 1 (3.8) | 1 (33.3) | 1 (3.8) | 1<br>(33.3) |
| Non-rebreather<br>mask | 0 (0.0) | 1<br>(33.3) | 0 (0.0) | 1<br>(33.3) | 0 (0.0) | 1 (33.3) | 0 (0.0) | 1 (33.3) | 0 (0.0) | 1<br>(33.3) |
| Oropharyngeal<br>airway (OPA) | 1<br>((3.8) | 1<br>(33.3) | 1 (3.8) | 1<br>(33.3) | 1 (3.8) | 1 (33.3) | 1 (3.8) | 1 (33.3) | 1 (3.8) | 1<br>(33.3) |
| Oxygen cylinder | 1 (3.8) | 2<br>(66.7) | 1 (3.8) | 2<br>(66.7) | 1 (3.8) | 2 (66.7) | 1 (3.8) | 2 (66.7) | 1 (3.8) | 2<br>(66.7) |
| Simple oxygen<br>face mask | 0 (0.0) | 1<br>(33.3) | 0 (0.0) | 1<br>(33.3) | 0 (0.0) | 1 (33.3) | 0 (0.0) | 1 (33.3) | 0 (0.0) | 1<br>(33.3) |

\*Combined; tablets and injections, I.V = Intravenous

### Average number of stockouts and longest stockout period [days]

Across the five years, dispensaries had more stockouts between 2019 and 2021; the highest observed stockout rate in dispensaries was 53.8% for chlorphenamine tablets in 2020. The health centers experienced a high stockout rate between 2021 and 2022, with the highest observed stockout rate being 66.7% for hydrocortisone injection in 2021. The corticosteroids, antihistamines, analgesics, and infusion fluid had a high number of stockouts. Among the antidotes for animal-related injury, the snake antivenom experienced a low stockout rate at dispensaries throughout the study period, with one facility (3.8%) reporting stockouts annually. However, stockout durations were substantial, ranging from 39.7 to 104 days, with the longest stockout recorded in 2022 (104 days). At health centers, one-third (33.3%) of facilities experienced stockouts each year, with stockout durations ranging from 30 to 60 days. Adrenaline injection showed relatively high stockout frequencies at dispensaries, affecting between 26.9% and 34.6% of facilities during the study period. The longest stockout duration increased from 36.8 days in 2019 to a peak of 103.6 days in 2021 before declining to 76.4 days in 2023. In contrast, health centers reported only one stockout event in 2019 (33.3%), with no stockouts recorded from 2020 to 2023.

The longest stockout period was observed in dispensaries, 218 days for IV giving set in 2019, while 102 days for dexamethasone injection in the health centers in 2020 (Table 2).

**Table 2.**
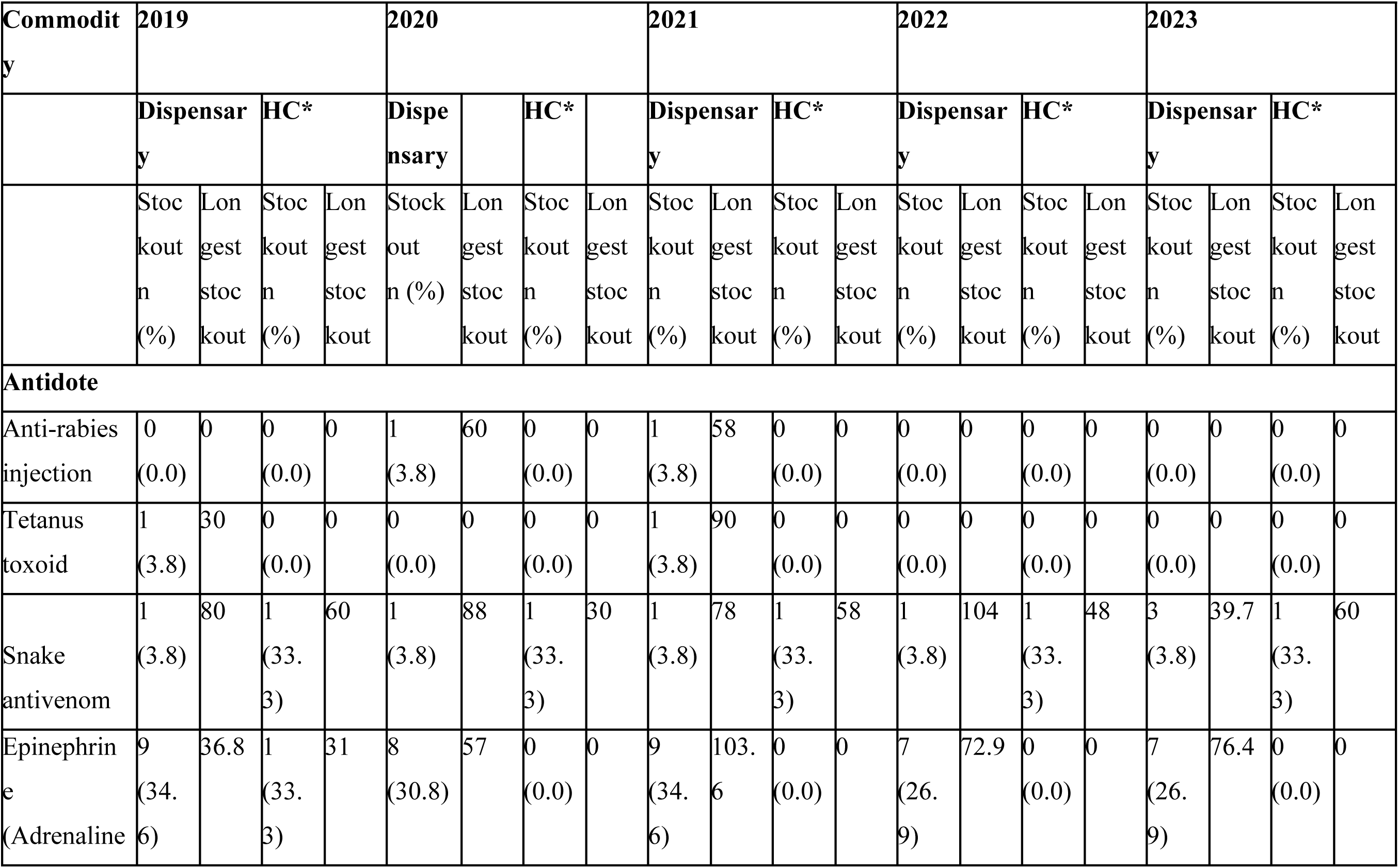
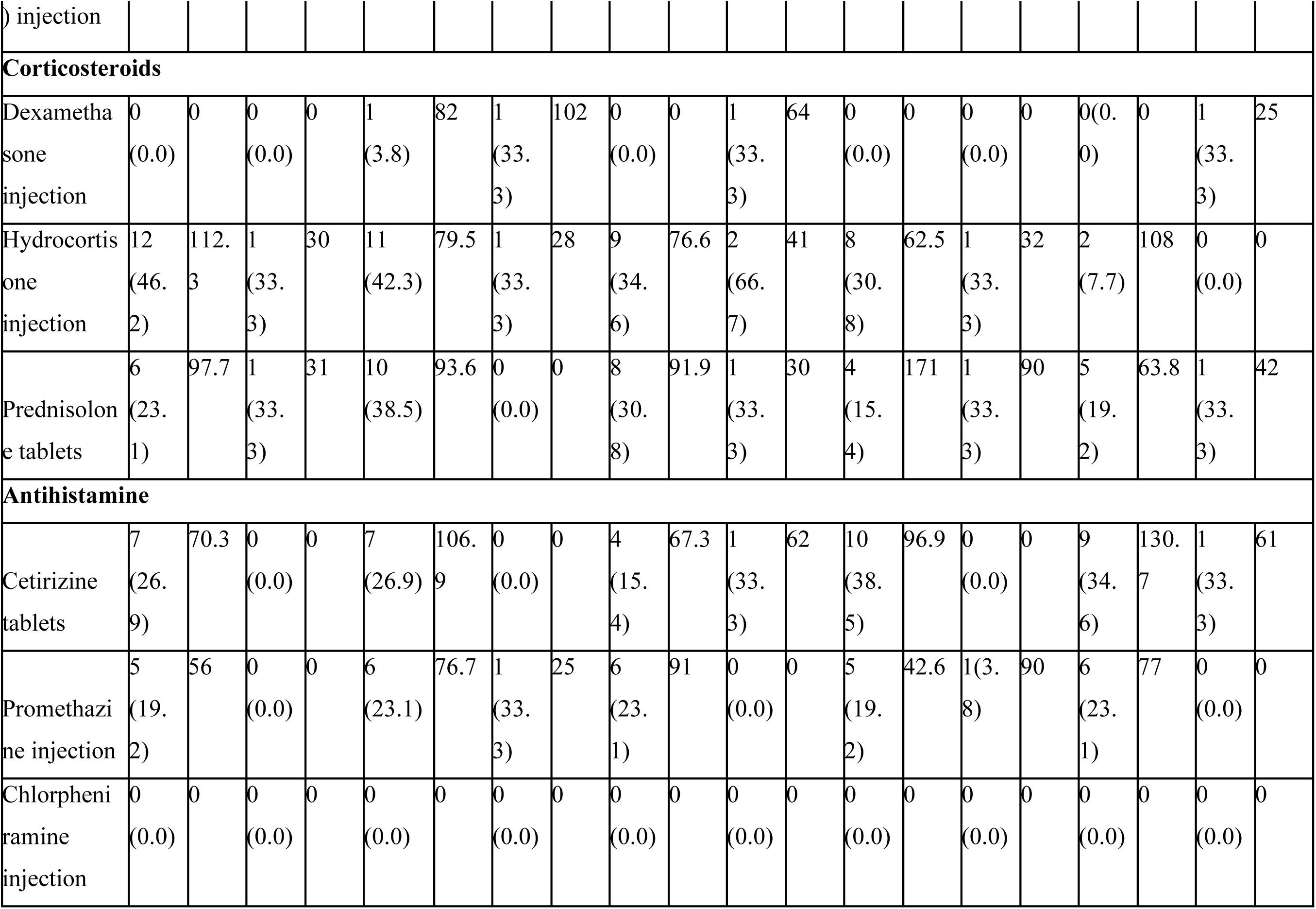
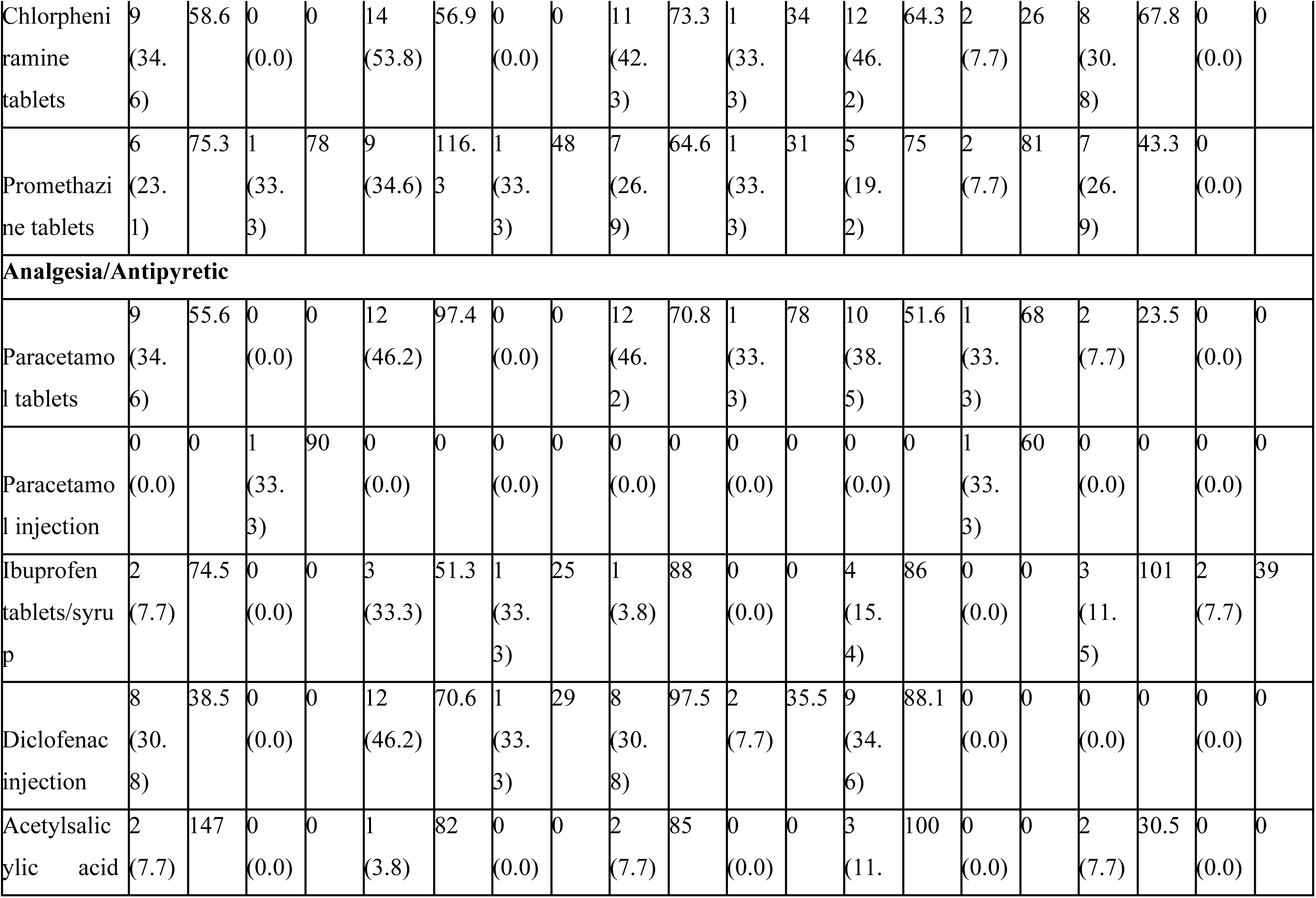
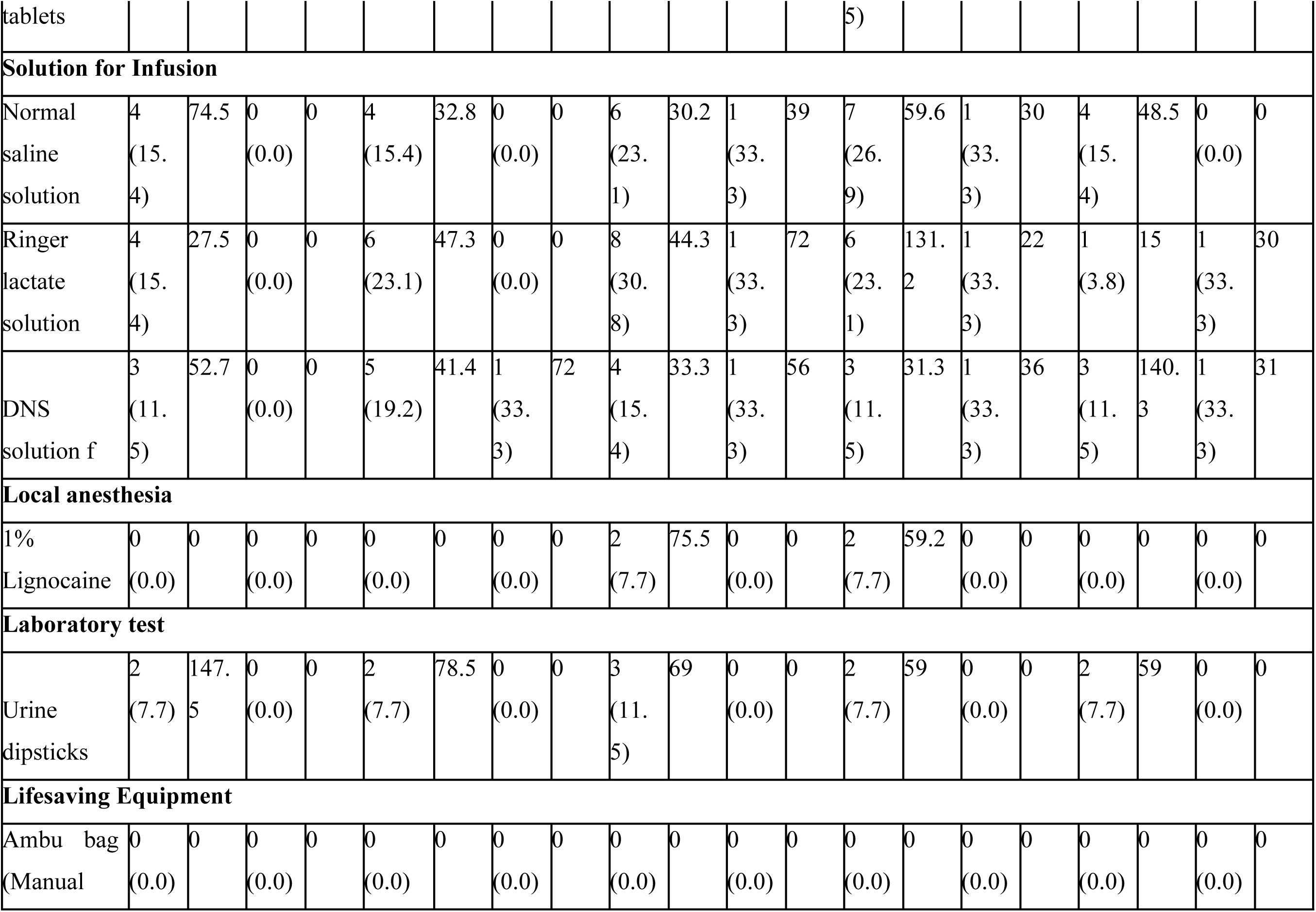
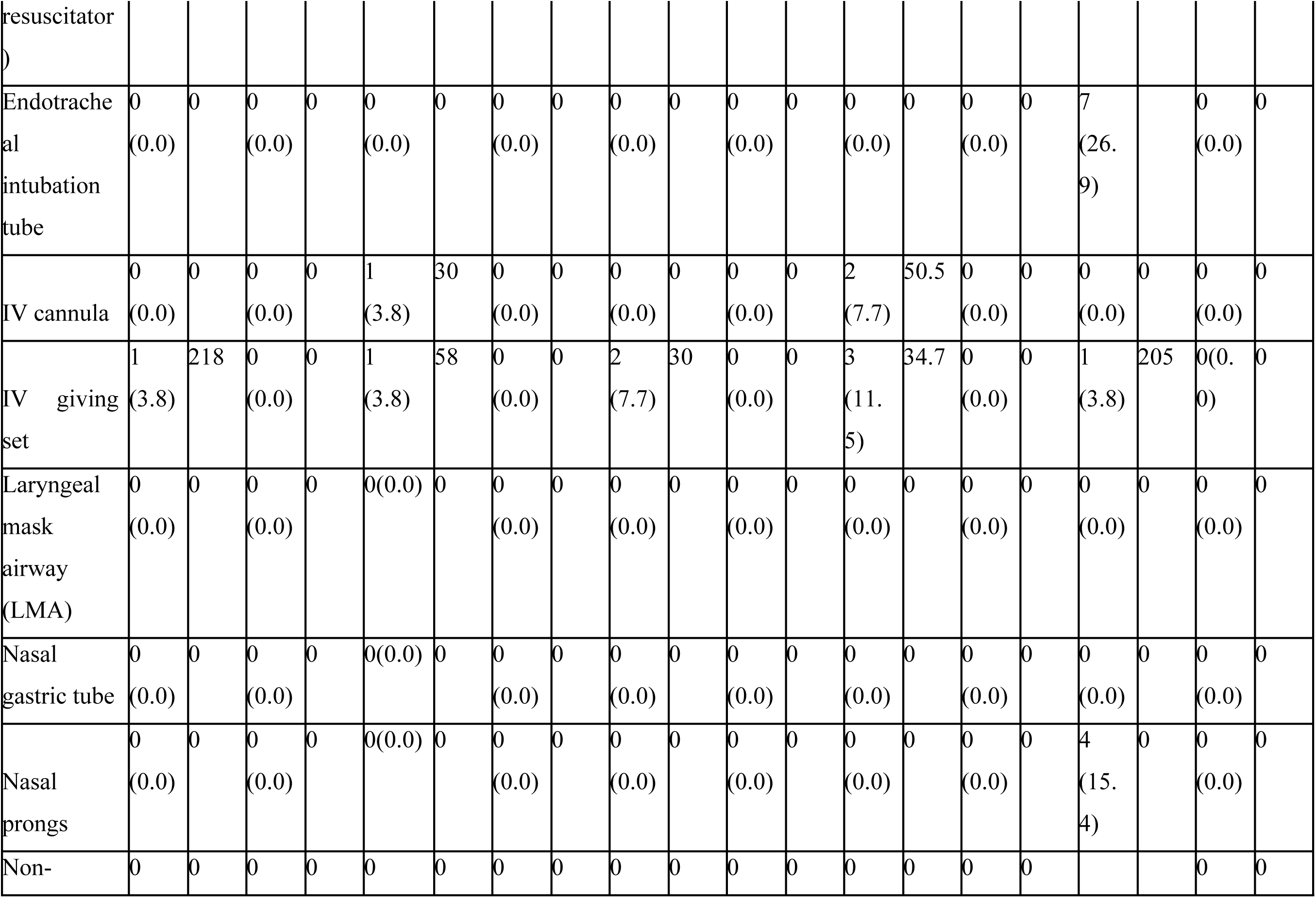
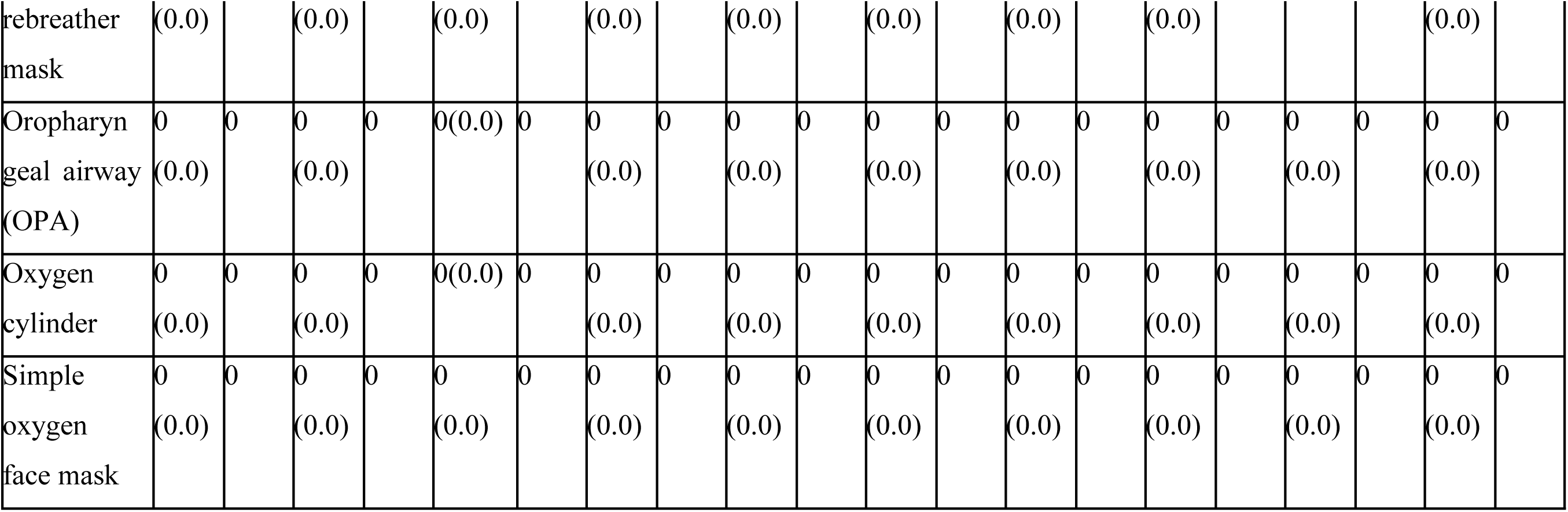
Average stockout rate and average longest stockout period (days) of animal-related injury commodities in visited Public Health Facilities in Mkinga District (n=29).

### Affordability of animal-related injuries commodities

Assuming all wages (100%) are used to purchase the medicines, most animal-related commodities were considered affordable for the lowest-paid government worker (LPGW), as it would take less than a day to purchase a full course of treatment. However, the Anti-rabies injection and Snake antivenom were not affordable, as it takes > 30 days for anti-rabies and > 300 days for snake antivenom. Assuming 40% of disposable income to purchase medicines, only epinephrine (adrenaline) injection, prednisolone, cetirizine, chlorpheniramine, promethazine, paracetamol, and ibuprofen tablets, and promethazine and chlorpheniramine injection were considered affordable. Using the affordability threshold of 7% of daily disposable income for healthcare expenditure, almost all assessed animal-related commodities were classified as unaffordable. The least affordable commodities were antidotes, for which the cost of a full course of treatment ranged from the equivalent of 11.1 to 18,571.0 days’ disposable income (Table 3).

**Table 3.** Affordability of animal-related injury commodities in visited Public Health Facilities in Mkinga District (n=29).

| Commodity | Treatment regimen unit | Median Price of Medicine and Medical Supplies (TZS) ~ USD per unit | Treatment duration per STG/NEML (days) | Affordability for a low government wage per day to buy a treatment course, assuming disposable income |  |  |
| --- | --- | --- | --- | --- | --- | --- |
|  |  |  |  | 100% | 40% | 7% |
| **Antidote** |  |  |  |  |  |  |
| Anti-rabies injection | 4 vials | (25,000) ~ 9.67 | 4 days (1 vial each day) | 32.5 | 81.2 | 464.3 |
| Tetanus toxoid | 1 vial | (2,400) ~ 0.93 | 1 day | 0.8 | 1.9 | 11.1 |
| Snake antivenom | Mild degree of envenomation –5 vials | (200,000) ~ 77.37 | 1 day | 325.0 | 812.5 | 4642.7 |
|  | Moderate degree of envenomation –10 vials |  |  | 650.0 | 1625.0 | 9285.5 |
|  | Severe degree of envenomation –20 vials |  |  | 1300.0 | 3250.0 | 18571.0 |
| Epinephrine | 1 vial | (1,100) ~ 0.43 | 1 day | 0.4 | 0.9 | 5.1 |
| (Adrenaline) injection |  |  |  |  |  |  |
| **Corticosteroids** |  |  |  |  |  |  |
| Dexamethasone injection | 1 vial | (1,200) ~ 0.46 | 1 day | 0.4 | 1.0 | 5.6 |
| Hydrocortisone injection | 1 vial | (1,300) ~ 0.50 | 1 day | 0.4 | 1.1 | 6.0 |
| Prednisolone tablets | 20 tablets | (50) ~ 0.02 | 5 days | 0.3 | 0.8 | 4.6 |
| **Antihistamine** |  |  |  |  |  |  |
| Cetirizine tablets | 3 tablets | (50) ~ 0.02 | 3 days | 0.05 | 0.1 | 0.7 |
| Promethazine injection | 1 vial | (1,000) ~ 0.39 | 1 day | 0.3 | 0.8 | 4.6 |
| Chlorpheniramine injection | 1 vial | (20) ~ 0.007 | 1 day | 0.01 | 0.01 | 0.1 |
| Chlorpheniramine tablets | 9 tablets | (50) ~ 0.02 | 3 days | 0.1 | 0.4 | 2.1 |
| Promethazine tablets | 3 tablets | (50) ~ 0.02 | 3 days | 0.05 | 0.1 | 0.7 |
| **Analgesia/Antipyretic** |  |  |  |  |  |  |
| Paracetamol tablets | 18 tablets | (31) ~ 0.01 | 3 days | 0.2 | 0.5 | 2.6 |
| Paracetamol injection | 9 bottles | (5,000) ~ 1.93 | 3 days | 14.6 | 36.6 | 208.9 |
| Ibuprofen tablets/syrup | 18 tablets | (50) ~ 0.02 | 3 days | 0.3 | 0.7 | 4.2 |
| Diclofenac injection | 9 vials | (800) ~ 0.31 | 3 days | 2.4 | 5.8 | 33.4 |
| Acetylsalicylic acid | 54 tablets | (100) ~ 0.04 | 3 days | 1.8 | 4.4 | 25.1 |
| tablets |  |  |  |  |  |  |
| **Solution for Infusion** |  |  |  |  |  |  |
| DNS solution for infusion | 1 bottle | (1500) ~ 0.58 | 1 day | 0.5 | 1.2 | 7.0 |
| Normal saline solution for infusion | 1 bottle | (1500) ~ 0.58 | 1 day | 0.5 | 1.2 | 7.0 |
| Ringer's lactate solution for infusion | 1 bottle | (1500) ~ 0.58 | 1 day | 0.5 | 1.2 | 7.0 |
| **Local anesthesia** |  |  |  |  |  |  |
| 1% Lignocaine solution for injection | 1 vial | (2000) ~ 0.77 | 1 day | 0.6 | 1.6 | 9.3 |
| **Laboratory test** |  |  |  |  |  |  |
| Urine dipsticks | 1 strip | (2000) ~ 0.77 | 1 day | 0.6 | 1.6 | 9.3 |
| **Lifesaving Equipment** |  |  |  |  |  |  |
| Ambu bag (Manual resuscitator) | 1 Ambu bag | (500) ~ 0.19 | 1 day | 0.2 | 0.4 | 2.3 |
| IV cannula | 1 cannula | (500) ~ 0.19 | 1 day | 0.2 | 0.4 | 2.3 |
| IV giving set | 1 set | (500) ~ 0.19 | 1 day | 0.2 | 0.4 | 2.3 |
| Nasal gastric tube (NGT) | 1 NGT | (1000) ~ 0.39 | 1 day | 0.3 | 0.8 | 4.6 |

## Discussion

This study assessed the availability, stockouts, and affordability of essential health commodities for managing animal-related injuries in public PHC facilities in Tanzania from 2019 to 2023. Overall, most of the 33 assessed commodities in dispensaries and health centers were available below the WHO target of > 80%. Furthermore, anti-rabies biologicals and snake antivenoms remained critically below. Adrenaline and dexamethasone injections, urine dipsticks, oxygen cylinders, simple oxygen face masks, and nasogastric tubes were also in low supply. Despite being at low availability, the majority of commodities experienced frequent stockouts. The highest stockout rates reached 53.8% in dispensaries and 66.7% in health centers, with some commodities remaining unavailable for over 200 days. Snake antivenom was out of stock for 104 days in dispensaries in 2022, while health centers recorded antivenom stockouts of 58 and 60 days in 2021 and 2023, respectively. Most commodities were affordable when assessed against a full daily wage, except the anti-rabies vaccine and snake antivenom. However, using the more realistic threshold of 7% of daily income for healthcare expenditure, nearly all commodities were unaffordable, with snake antivenom being the least affordable.

Snake antivenom and anti-rabies biologicals are classified by the WHO as essential, life-saving medicines and should be available across all levels of healthcare [6]. Consistent with findings from previous studies, this study found low availability of snake antivenom and anti-rabies in PHC facilities [9]. Although snake antivenom products became universally available in health centers from 2022, their availability in dispensaries remained below the recommended level. Previous studies from SSA have reported snake antivenom availability ranging from 4% to 47% in public hospitals and 10% to 20% in private health facilities [9–11]. Even where these commodities are available, prolonged stockouts remain common, particularly in rural settings. Similar findings were observed in this study, where some animal-related injury commodities experienced stockouts lasting up to 200 days. Such interruptions compromise the readiness of primary healthcare facilities to manage animal-related injuries [20]. This, in turn, delays timely access to essential treatment and may increase preventable morbidity and mortality.

The limited availability of these commodities is partly attributed to the centralized procurement systems commonly used across SSA countries [9,21]. In many settings, forecasting and procurement decisions rely largely on historical consumption data rather than epidemiological burden [22]. This challenge is particularly pronounced for animal-related injuries, where the true burden of disease and geographical hotspots remain poorly surveyed and documented. Consequently, forecasting, inventory management, and distribution of animal-related injury commodities are often suboptimal. Additional factors reported in the literature include limited numbers of manufacturers and suppliers, procurement and tendering challenges, competing healthcare priorities, and inadequate supply chain management skills among healthcare workers [9,23]. Therefore, to strengthen service availability and ensure PHC facilities are adequately prepared to provide timely and effective care, measures such as community-based animal-related injury surveillance and engagement of traditional healers in case reporting should be implemented. These approaches can improve estimation of the true epidemiological burden and identification of geographical hotspots, thereby informing data-driven forecasting, procurement, allocation, and distribution of essential commodities for managing animal-related injuries.

This study did not assess the quantities of commodities available at health facilities. This is an important consideration because management of animal-related injuries often requires multiple doses or vials of treatment, and the mere presence of a commodity may not be sufficient to meet patient needs [18]. Besides, the study did not evaluate the effectiveness of available snake antivenoms against medically important snake species found in Tanzania. This is particularly relevant given reports from Kenya showing that some commercially available antivenoms demonstrated limited effectiveness in clinical trials, while studies from Ghana have documented deaths among snakebite victims despite the timely administration of antivenom – this is contributed by the vast species of snake in the African continent [11,24]. Future studies should assess not only the availability but also the quantities of animal-related injury health commodities in healthcare facilities. In addition, the effectiveness of available snake antivenoms and anti-rabies vaccines should be evaluated. This is particularly important for snake antivenoms, as their clinical effectiveness varies according to the snake species responsible for envenomation and the extent to which the antivenom is matched to locally prevalent venomous snakes.

This study also found that animal-related injury commodities were largely unaffordable for victims and their caregivers. These findings are consistent with previous studies conducted in SSA, which have highlighted the substantial financial burden associated with accessing treatment for snakebites and rabies exposures [9–11]. In Tanzania, the cost of a single vial of snake antivenom ranges from USD 60 to 80, which falls within the broader African range of approximately USD 4 to 200 per vial [9]. However, snakebite victims often require multiple vials of antivenom, sometimes up to eight or more, in addition to supportive treatments and hospitalization [18]. Consequently, the total cost of snakebite management in Africa has been estimated to range from USD 55 to 6,250 per patient, placing treatment beyond the reach of many households.

The poor affordability of animal-related injury commodities is partly attributable to the low prioritization of these conditions within healthcare systems in many SSA countries [9]. As NTDs, snakebite envenoming and other animal-related injuries often receive less attention than major public health programs such as HIV/AIDS, tuberculosis, and reproductive, maternal, newborn, and child health services [12]. As a result, these conditions are allocated a limited or no dedicated budget, attract relatively little donor support, and are often excluded from medicine price-subsidization schemes and exemption policies.

Furthermore, the costs of key treatments, including snake antivenom and anti-rabies biologicals, are frequently not fully covered by health insurance schemes in many SSA countries. Consequently, victims and their caregivers are required to pay for treatment out-of-pocket. Such out-of-pocket healthcare expenditures have been widely recognized as a major barrier to accessing and utilizing essential healthcare services in low- and middle-income countries [25,26]. High treatment costs may delay care-seeking, lead to treatment discontinuation, and expose affected households to catastrophic health expenditures, thereby exacerbating poverty and health inequities. Immediate efforts are needed to ensure that life-saving medicines for animal-related injuries are covered under commonly used health insurance schemes while promoting enrolment in available insurance programs. Where feasible, countries in SSA should introduce subsidies for snake antivenoms and anti-rabies vaccines to improve affordability and access. Existing exemption policies should also be strengthened to ensure that vulnerable and disadvantaged populations receive these essential medicines free of charge. Collectively, these measures would enhance the availability of essential commodities, improve the readiness of PHC facilities, which provide care for the majority of the population, and accelerate progress toward achieving UHC.

Affordability in this study was assessed using the LPGW method, as recommended by the WHO/ HAI medicine pricing methodology. While this approach is widely used, it has several limitations. It does not account for household dependents, additional healthcare costs such as consultation fees, diagnostic tests, transportation, and food expenses incurred during treatment. Furthermore, it assumes formal employment and does not adequately reflect the financial realities of unemployed individuals or those working in the informal sector, who constitute a substantial proportion of the population in SSA. Therefore, the affordability estimates presented in this study should be interpreted with caution, as they may underestimate the true financial burden faced by patients of animal-related injury and their caregivers.

## Conclusions

This study demonstrates substantial health system gaps in the availability, continuity of supply, and affordability of essential commodities for managing animal-related injuries in rural Tanzania. Anti-rabies biologicals and snake antivenom consistently failed to meet the WHO-recommended availability threshold, particularly in dispensaries, while deficiencies in emergency medicines, diagnostic supplies, and supportive care commodities further limited the capacity of PHC facilities to deliver timely, life-saving care. Frequent and prolonged stockouts, together with the poor affordability of most commodities, expose rural populations to preventable morbidity, mortality, and catastrophic out-of-pocket expenditure. Addressing these gaps requires coordinated policy and health system reforms that strengthen surveillance of animal-related injuries, demand forecasting, procurement, financing, and supply chain management. Expanding financial risk protection through health insurance and targeted subsidies for high-cost commodities, alongside equitable distribution of essential supplies to PHC facilities, will be critical to improving emergency preparedness, reducing inequities in access to care, and advancing UHC in Tanzania and other developing countries.

## Acknowledgments

The authors sincerely thank Ms. Lucia Mgaya from the Mkinga District Council for her invaluable support in facilitating the implementation of this study. We are also grateful to the University of Rwanda through the East African Community Regional Centre of Excellence for Vaccines, Immunization and Health Supply Chain Management [EAC RCE-VIHSCM] Research Grants for providing financial support for this study. We further acknowledge the contributions of all research assistants who supported data collection.

The authors used ChatGPT [OpenAI] solely to assist with language editing and improving the clarity and readability of the manuscript. All scientific content, interpretation of findings, and final editorial decisions were made by the authors, who take full responsibility for the manuscript.

## Authors’ contributions

Manase Kilonzi^1*^, Joseph Matobo Thobias^2^, Gimbo Hyuha^3^, Paul Makoye Malaba^1^, Beatrice Aiko^1^, George Kiwango^4^, Alphonce Ignace Marealle^1,^ Nathanael Sirili^3^

**Conceptualization:** Manase Kilonzi, Beatrice Aiko, Paul Makoye Malaba, Gimbo Hyuha, Nathanael Sirili.

**Data curation:** Manase Kilonzi, Joseph Matobo Thobias, Gimbo Hyuha, Paul Makoye Malaba, Beatrice Aiko, George Kiwango, Alphonce Ignace, Marealle, Nathanael Sirili.

**Formal analysis:** Manase Kilonzi, Joseph Matobo Thobias, Gimbo Hyuha, George Kiwango.

**Funding acquisition:** Manase Kilonzi, Beatrice Aiko, Paul Makoye Malaba, Gimbo Hyuha, Nathanael Sirili.

**Investigation:** Manase Kilonzi, Joseph Matobo Thobias, Paul Makoye Malaba, Beatrice Aiko, George Kiwango.

**Methodology:** Manase Kilonzi, Gimbo Hyuha, Paul Makoye Malaba, Beatrice Aiko, George Kiwango, Alphonce Ignace Marealle, Nathanael Sirili.

**Project administration:** Manase Kilonzi, Nathanael Sirili, Gimbo Hyuha, Paul Makoye Malaba, Beatrice Aiko, Joseph Matobo Thobias, George Kiwango, Alphonce Ignace Marealle.

**Resources:** Manase Kilonzi, Beatrice Aiko, Paul Makoye Malaba, Gimbo Hyuha, Nathanael Sirili.

**Software:** Manase Kilonzi, Joseph Matobo Thobias.

**Supervision:** Nathanael Sirili, Manase Kilonzi, Alphonce Ignace Marealle.

**Validation:** Manase Kilonzi, Joseph Matobo Thobias, Gimbo Hyuha, Paul Makoye Malaba, Beatrice Aiko, George Kiwango, Alphonce Ignace, Marealle, Nathanael Sirili.

**Visualization:** Manase Kilonzi, Joseph Matobo Thobias.

**Writing – original draft preparation:** Joseph Matobo Thobias, Manase Kilonzi, Gimbo Hyuha.

**Writing – review & editing:** Manase Kilonzi, Joseph Matobo Thobias, Gimbo Hyuha, Paul Makoye Malaba, Beatrice Aiko, George Kiwango, Alphonce Ignace, Marealle, Nathanael Sirili.

## Availability of data and materials

The datasets used and/or analyzed during the current study are available from the corresponding author at a reasonable request.

